# Short-term forecasts and long-term mitigation evaluations for the COVID-19 epidemic in Hubei Province, China

**DOI:** 10.1101/2020.03.27.20045625

**Authors:** Qihui Yang, Chunlin Yi, Aram Vajdi, Lee W. Cohnstaedt, Hongyu Wu, Xiaolong Guo, Caterina M. Scoglio

## Abstract

As an emerging infectious disease, the 2019 coronavirus disease (COVID-19) has developed into a global pandemic. During the initial spreading of the virus in China, we demonstrated the ensemble Kalman filter performed well as a short-term predictor of the daily cases reported in Wuhan City. Second, we used an individual-level network-based model to reconstruct the epidemic dynamics in Hubei Province and examine the effectiveness of non-pharmaceutical interventions on the epidemic spreading with various scenarios. Our simulation results show that without continued control measures, the epidemic in Hubei Province could have become persistent. Only by continuing to decrease the infection rate through 1) protective measures and 2) social distancing can the actual epidemic trajectory that happened in Hubei Province be reconstructed in simulation. Finally, we simulate the COVID-19 transmission with non-Markovian processes and show how these models produce different epidemic trajectories, compared to those obtained with Markov processes. Since recent studies show that COVID-19 epidemiological parameters do not follow exponential distributions leading to Markov processes, future works need to focus on non-Markovian models to better capture the COVID-19 spreading trajectories. In addition, shortening the infectious period via early case identification and isolation can slow the epidemic spreading significantly.

## 1. Introduction

The 2019 novel coronavirus disease (COVID-19) emerged in Wuhan City, Hubei Province of China, in the late fall of 2019 (Chen et al., 2020). The highly contagious virus outbreak during the Chinese Spring festival period made containment an unprecedented challenge. The risks of human-to-human transmission, long persistence on surfaces, and asymptomatic infections increased spread and made case detection difficult (Chan et al., 2020; Rothe et al., 2020). Increasing case reports saturated hospitals in Hubei Province, overwhelming medical resources. Despite the high case reports, the true infection rate was likely underreported, as infected individuals with only mild symptoms might have recovered without being detected (Nishiura et al., 2020; Sanche et al., 2020).

To stop the viral spread, a series of strict control measures have been implemented in mainland China, including the enforcement of the tourism ban, suspension of the school, and extension of the Spring Festival holiday. From January 23rd to April 8^th^, 2020, the Chinese government implemented a metropolitan-wide quarantine of Wuhan and nearby cities, suspending all public transportation and advising residents to stay at home (Du et al., 2020; J.T. Wu, Leung, & Leung, 2020). Since February 8^th^, a door-to-door check was started in Wuhan to take all confirmed and suspected people into medical care, which ended at around February 19^th^ (Zhao et al., 2020). Since February 10^th^, Wuhan City started to close the community.

Understanding the impact of non-pharmaceutical interventions on the epidemic dynamics is crucial for decision-makers to take proper actions to contain the COVID-19. Since the beginning of the outbreak, scientists have been actively retrieving the epidemiological characteristics such as the time from infection to illness onset (incubation period) and the time from illness onset to hospital admission or isolation (infectious period) (Chen et al., 2020; Guan et al., 2020; Huang et al., 2020). Researchers have used the reported cases during the early stage of the epidemic and predicted the epidemic trajectory, the basic reproductive number *R*_0_ from which ranges from 2.2 to 6.6 (Read, Bridgen, Cummings, Ho, & Jewell, 2020; Sanche et al., 2020; Zhao et al., 2020). Several studies explored the effects of the quarantine of Wuhan (X. Li, Zhao, & Sun, 2020; Shen, Peng, Guo, Xiao, & Zhang, 2020), travel restrictions (Chinazzi et al., 2020), and other non-pharmaceutical intervention strategies (Lai et al., 2020; Zhu et al., 2020) on the future transmission dynamics at different geographical scales.

China’s aggressive and prompt measures have led to a dramatic decrease in the number of new cases. Those measures saved many lives, although at a high economic cost. As of March 18^th^, 2020, the cumulative number of confirmed cases in China was 81,186. Compared to the previous day, the number of new cases was only 70, most of which were imported from outside the country (T. Wu, Ge, Yu, & Hu, 2020). In the meantime, COVID-19 spread further around the world, and parts of Italy, South Korea, the United States, and Iran struggled to contain the epidemic.

In this work, we aim to assess the capability of the ensemble Kalman filter as a good short-term predictor, and test the effectiveness of non-pharmaceutical interventions on the epidemic spreading. First, we provide real-time assessments and forecasts of the case reported in Wuhan City based on the ensemble Kalman filter. Second, we build an individual-level based network model and perform stochastic simulations to reconstruct the epidemics in Hubei Province at its early stage and examine the epidemic dynamics under different scenarios. We consider four scenarios: keeping the early-stage trend without any mitigation, reducing the average node through social distancing, reducing the infection rate by adopting protective measures, and decreasing the infection rate through both social distancing and protective measures. Third, we simulate the epidemic spreading in Wuhan City incorporating non-Markovian processes, i.e., non-exponential parameter distributions, and compare the predicted epidemic trajectories obtained from Markov processes.

Mathematical modeling can forecast the size and timing of epidemics, so resource allocations and decision-making can be optimized to maximize intervention strategies. Since most current network-based models are based on Markov processes, this work is probably one of the first to simulate the COVID-19 spreading based on non-Markovian processes. The findings of this work can be a reference for policy recommendations regarding COVID-19 in other countries.

## 2. Model

In this section, we first introduce the ensemble Kalman filtering method used in the short-term prediction. Then we illustrate epidemic models based on Markov processes and non-Markovian processes built in the long-term analysis.

To describe the epidemiology of COVID-19 pneumonia, we use the Susceptible-Exposed-Infectious-Removed (SEIR) compartmental model, which has been frequently used in the study of the COVID-19 (X. Li et al., 2020; Read et al., 2020; J. T. Wu et al., 2020; Zhao et al., 2020). Each node is either in one of the compartments susceptible, exposed, infectious, or removed. Susceptible nodes refer to people free of the virus and can become infected due to contact with infectious nodes. Exposed nodes have been infected by the disease, but have not yet shown symptoms or become infectious. Removed nodes represent those individuals who are recovered, isolated, hospitalized, or dead due to the disease.

Right after the COVID-19 outbreak in China, many studies have estimated the epidemiological parameters, which are essential to calibrate the epidemic models. However, we noticed that there exists a variety regarding the epidemiological parameters. Based on 1099 patients in 31 provincial municipalities through January 29^th^, 2020, Guan et al. (2020) estimated that the median incubation period was 4.0 days (interquartile range 2 to 7 days). Based on a smaller sample size of 425 patients, Q. Li et al. (2020) estimated the mean incubation period to be 5.2 days (95% confidence interval, 4.1 to 7.0 days). Du et al. (2020) considered the delay from symptom onset to case detection as 4–5 days. Considering the possibility of asymptomatic infections, we adopt a relatively short mean incubation (latent) period as 3 days and mean infectious period as 4 days in the short-term prediction and long-term analysis based on Markovian processes (Lin et al., 2020).

### 2.1 Short-term prediction based on Kalman filtering

Short-term prediction is provided for the COVID-19 spreading in Wuhan city using the Kalman filtering, which consists of model state estimation at each observation time based only on the observation up to the present. Considering that there is an unknown time-varying parameter *β* in the SEIR model which cannot be dealt with by widely used classic Kalman filter (Kalman, 1960) and extended Kalman filter (Jazwinski, 2007), a dual state-parameter estimation filtering method-ensemble Kalman filter (EnKF) is used for our study (Jensen, 2007; Moradkhani, Sorooshian, Gupta, & Houser, 2005). EnKF is based on Monte Carlo, where the approximation of forecast (a priori) state error covariance matrix is made by propagating an ensemble of model states using the updated states (ensemble members) from the previous time step.

We use a generalized network, in which every individual is connected with a possibility. At any discrete time, every individual has to be in one of the four compartments in the SEIR compartmental model. The equations for the state transitions are shown as follows.

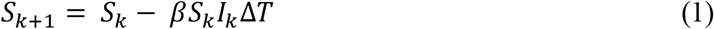

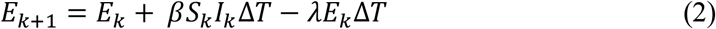

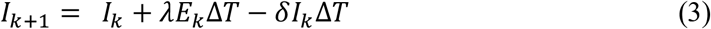

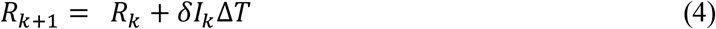

Where *β* is the infection rate; *λ* is the transition rate from the exposed compartment to the infectious compartment; *δ* denotes the removal rate. Δ*T* = 1 day and Δ*T* = 0, 1, 2, 3 is the *k*^th^ time step. We assume that the infection process of each infectious individual is Poisson and statistically independent, and that the removing process for every infected individual is also Poisson. The infection rate *β* quantifies the probability that each susceptible individual can be infected by each infectious individual in a time unit. The rate *λ* quantifies the average portion of the exposed population that will become infectious in a time unit. The removal rate *δ* quantifies the average portion of the infectious population that will clear the infection in a time unit. Considering that the population flow, prevention measures, and self-quarantine could affect the infection rate, we take the infection rate *β* as a time-varying parameter. The other two rates *λ* and *δ* are set as constants 1/3 day^-1^ and 1/4 day^-1^ (Lin et al., 2020).

Let the state variables (SEIR) defined as *x*_*k*_ ∈ *R*^4^, the time-varying parameter *β*_*k*_ ∈ *R* Consider the following problem

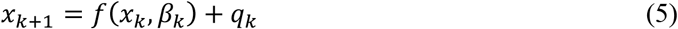

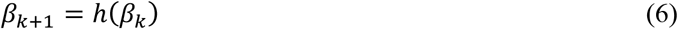

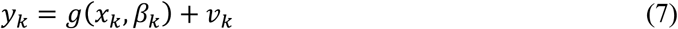

where *q*_*k*_ and *v*_*k*_ are model and observation noise which are assumed to be normally distributed and with zero mean and the covariance known.

As the true states are generally unknown, we use the ensemble mean as the best estimate of the true state. The error covariance matrices for the predicted and filtered ensemble are given by

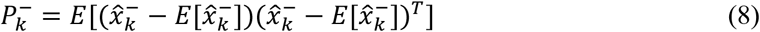

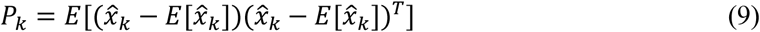

where 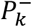 is the error covariance matrix associated with the forecasted estimate (prior), *P*_*k*_ is the one associated with the updated estimate (posterior), 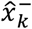 is the ensemble vector of the forecasted estimate from model equations.

In the ensemble presentation, the model states *x*_*k*_ and the time-varying parameter *β*_*k*_ can be put together in a matrix *A*_*k*_ ∈ *R*^5×*N*^, holding *N* ensemble member at time *k*.

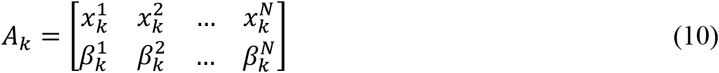

The ensemble covariance then can be presented as

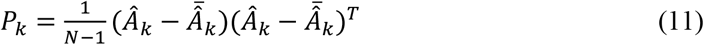

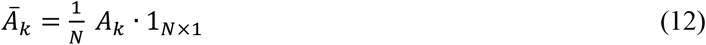

The measurement can also be formulated in an ensemble presentation

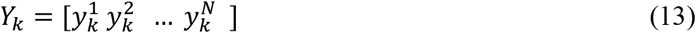

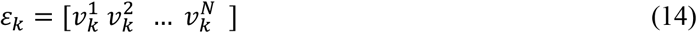

where 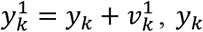 is the true measurement and 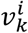 is the perturbation term. The measurement covariance matrix can be estimated as

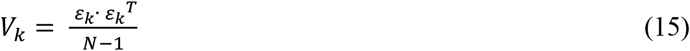

Specify to the situation we are dealing with, *A*_*k*_ = [*S*_*k*_ *E*_*k*_ *I*_*k*_ *R*_*k*_ *β*_*k*_]^*T*^ where 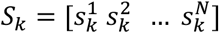 and it is the same with *E*_*k*_, *I*_*k*_, *R*_*k*_, and *β*_*k*_. Note that 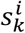 denotes the *i*^th^ sample in step *k*. The measurement is the total number of confirmed cases reported by the National Health Commission of China, which is compared with the number of removed population in our model. Therefore, *y*_*k*_ *= C* · [*x β*]^*T*^ where *C =* [0 0 0 1 0].

As the criteria for counting the confirmed cases are changed on February 12^th^, 2020, there is a jump in the reported number of confirmed cases. Taking this into account, we separate the whole time period into period 1 (from January 26^th^ to February 11^th^) and period 2 (from February 12^th^ to February 27^th^), and run simulations in two steps. First, we run simulations for period 1 and estimate the infection rate *β*. Then, we take the estimated *β* on February 11^th^ and the reported number of cases on February 12^th^ as the initial *β* and initial removed population for period 2, respectively, and run simulations. The number of iterations is 250 in every predictive simulation.

### 2.2 Long-term analysis with the Markovian process

In this section, we apply the generalized modeling framework (GEMF) proposed by Sahneh, Scoglio, & Van Mieghem (2013) to build an individual-level network model. In GEMF, each node stays in a different state, and the joint state of all nodes follow a Markovian process. The node-level description of this Markovian process can be expressed as follows:

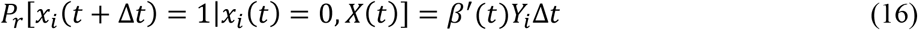

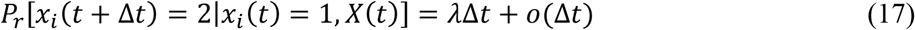

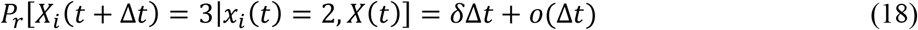

Where *X*(*t*) is the joint state of all individual nodes at time *t*; *x*_*i*_(*t*) = 0, 1, 2, 3 means that the state of node *i* at time *t* is in susceptible, exposed, infectious and removed compartment, respectively; *Y*_*i*_ is the number of infectious neighbors of node *i*; *β*′(*t*) is the rate for one node transitions from susceptible state to the exposed state; *λ* is the rate from the exposed state to the infectious state, and *δ* is the removal rate from infectious state to the removed state, which is the inverse of the infectious period.

In the network, nodes represent people and are divided to 17 groups to represent the 17 prefecture-level cities in Hubei Province (each group is called a city hereafter). Links are generated to reflect both the effects of person-to person contacts within a city and population flows between the cities on the spread of COVID-19. More specifically, we generate intra-city links for each city in two steps. First, we further divide nodes in each city into multiple communities to make it more realistic. Inside each community, links are generated based on the degree distribution given in Zhang et al. (2020) using the configuration model. Second, on average each node is connected to another node in the same city based on Erdős–Rényi model such that these communities have connections with each other.

Then, we generate inter-city links based on the number of people that traveled between the cities obtained from Baidu Migration data. For example, if the daily population flow from city *i* to city *j* is *K* number of people, directed links will be generated between *K* randomly selected nodes in city *i* to *Kd*_*t*_ nodes in city *j*, where *d*_*t*_ is the network average node degree at time *t*. More specifically, the average daily population movement from January 1^st^–22^nd^ is used to build *inter-city links before the Wuhan City lockdown* on January 23^rd^. Since the value of population flows between cities remain stable during the quarantine period, we use the average daily population movement from January 23^rd^–29^th^ to build *inter-city links after the Wuhan City lockdown*. Details about data collection can be found in Appendix.

In GEMF simulations, we consider the COVID-19 epidemic outbreak starting in Wuhan City from January 19^th^, because all confirmed cases reported in Hubei Province were in Wuhan City before January 19th, 2020. To reduce the computational cost, the population of each city and population flows among the cities are scaled by 198, which is the number of cumulative confirmed cases in Hubei Province up to January 19^th^. After scaling, the total number of nodes in the network *N* equals 298,838. In total, we generate three adjacency lists of the network topology, which are imported to GEMF and changed according to time during simulation. More specifically, the first adjacency list *A*_1_ corresponds to the network with *d*_*t*_=15 and inter-city links before the Wuhan City lockdown, and is used during simulation up to January 22^nd^. The second adjacency list *A*_2_ refers to the network with *d*_*t*_=15 and inter-city links after the Wuhan City lockdown, and is used during simulation up to February 9^th^. The third adjacency list *A*_3_ incorporates *d*_*t*_=3 and inter-city links after the Wuhan City lockdown, and is used during simulation on and after February 10^th^ when community closure was implemented in Wuhan city.

To fit the model, we take the number of confirmed cases as the number of people in the removed state. By keeping *λ* = 1/3 day^-1^ and *δ* = 1/4 day^-1^ unchanged, we estimate the infection rate *β*′ as a piecewise constant function of the time. The *β*′ function and time intervals are both tuned to minimize the error between the number of removed people from our simulations and the number of cumulative confirmed cases in Hubei Province (Health Commission of Hubei Province, 2020). The total period from January 19^th^ to April 15^th^ is separated into three phases (January 19^th^–January 27^th^, January 28^th^–February 9^th^, and after February 9^th^). During the first phase, Wuhan city lockdown happened, and hospitals were short of bed. During the second phase, Thunder God Mountain Hospital and Fire God Mountain Hospital were put into use, and door-to-door screening was implemented. In the third phase, community closure was enacted.

GEMF is available in MATLAB, R, Python, and C programming language (Sahneh, Vajdi, Shakeri, Fan, & Scoglio, 2017; Yang, Lu, Scoglio, de Jong, & Gruenbacher, 2018; Yi, Yang, & Scoglio, 2020). We performed extensive stochastic simulations by adapting the GEMF toolbox with MATLAB during the experiment. We track the number of nodes in each compartment and demonstrate the simulation results with confidence intervals. The number of simulation runs is 1500 in all scenarios.

### 2.3 Long-term analysis with the non-Markovian process

Since the outbreak of COVID-19, many studies have emerged to learn how this disease spreads. However, an overwhelming majority of the epidemic models have considered Markov processes, in which the transition between compartments follows an exponential distribution. To the best of the authors’ knowledge, very few works have been done incorporating non-Markovian processes to epidemic models with respect to COVID-19. In this section, we take Wuhan City as a study area and try to demonstrate how distributions of key epidemiological factors can influence the predicted trajectory of the disease spreading.

More specifically, we incorporate non-exponential interevent time distributions to the SEIR model using the modified Gillespie algorithm proposed by Boguñá, Lafuerza, Toral, & Serrano (2014). Transitions from the susceptible state to the exposed state is a Poisson process that follows the exponential distribution. Following lognormal distributions, the mean and median of the incubation period are estimated at 5.6 days and 5.0 days, respectively, while the infectious period is of mean 3.9 days and median 1.5 days (Linton et al., 2020), which is in line with Sanche et al. (2020). The number of simulation runs is 80.

## 3. Results and discussion

In this section, we provide results regarding the short-term analysis with the Kalman filter. From the long-term perspective, we first present results based on GEMF stochastic simulations and then compare the results based on non-Markovian processes and Markov processes.

### 3.1 Short-term analysis with the Kalman filter

The predictions of COVID-19 cases in Wuhan city from February 22^nd^ to March 1^st^ are conducted. The transition rates *λ* and *δ* are 1/3 day^-1^ and 1/4 day^-1^, respectively. The infection rate is estimated by EnKF. The model noise and measurement noise are tuned by modifying the approach proposed by R. Li et al. (2020).

Fig. 1 shows the distributions of the predicted numbers of COVID-19 cases for 1-day-ahead, 2-day-ahead, and 3-day-ahead. The solid black lines indicate the maximum and minimum prediction results on each day. The blue box presents the range from the lower quartile to the upper one. The red line inside the box indicates the median of the samples. We can see that most predictions converge to a small range, and that most medians are very close to the corresponding real data. Taking the median as our predicted number gives the relative error shown in Fig. 1(d). The mean absolute percentage error (MAPE) for 1-day-ahead prediction is 0.24%; the MAPE for 2-day-ahead prediction is 0.52%; the one for 3-day-ahead is 1.23%. The performance of 1-day-ahead prediction is the best and the one of 3-day-ahead is the worst. When the *n*-day-ahead prediction is conducted, the results from (*n*-1)-day-ahead is used, so the errors accumulate.

**Fig 1.**
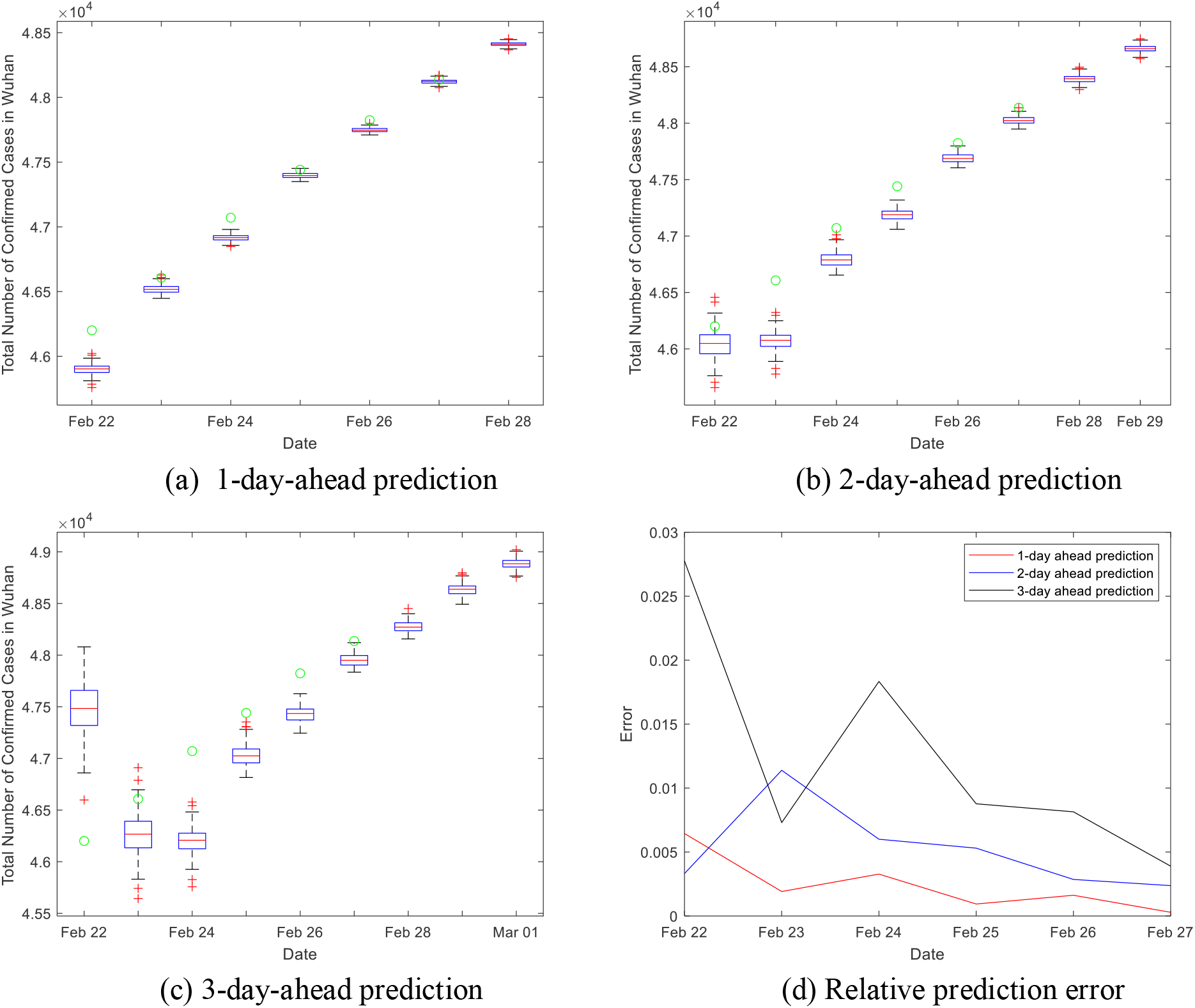
Distributions of the short-term prediction.

If we choose the average number of the samples as our predicted number of COVID-19 cases, the results and their relative errors are presented in Fig. 2. The MAPE for 1, 2, 3-day-ahead predictions are 0.24%, 0.52% and 1.23%, which are equal to the ones when the medians are predicted numbers.

**Fig 2.**
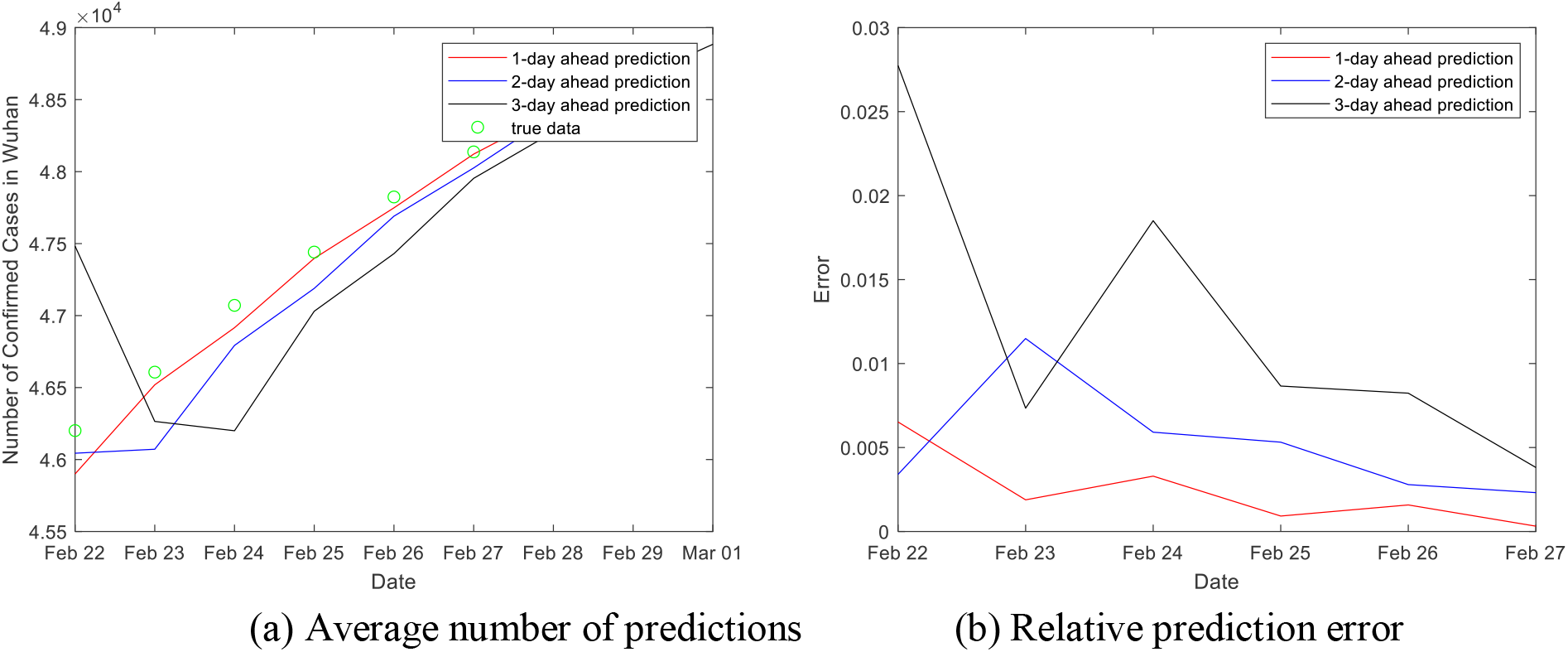
Average number as predictions and the error.

The self-quarantine policy was carried out on January 23^rd^ in Wuhan and a series of administrative actions were taken to prevent the disease from spreading. Taking the latent period into consideration, we started to estimate the infection rate *β* from January 26^th^. On February 12^th^, Wuhan authorities changed the case definition to include symptomatic patients in addition to tested ones. Therefore, we present estimations of *β* for period 1 (from January 26^th^ to February 11^th^) and period 2 (from February 12^th^ to February 27^th^), which are presented as the blue line and the red line in Fig. 3. The orange line in Fig. 3 presents the removal rate. From the results, we see that the infection rate *β* is less than the removal rate *δ* after February 10^th^, which means the epidemic had peaked in Wuhan.

**Fig 3.**
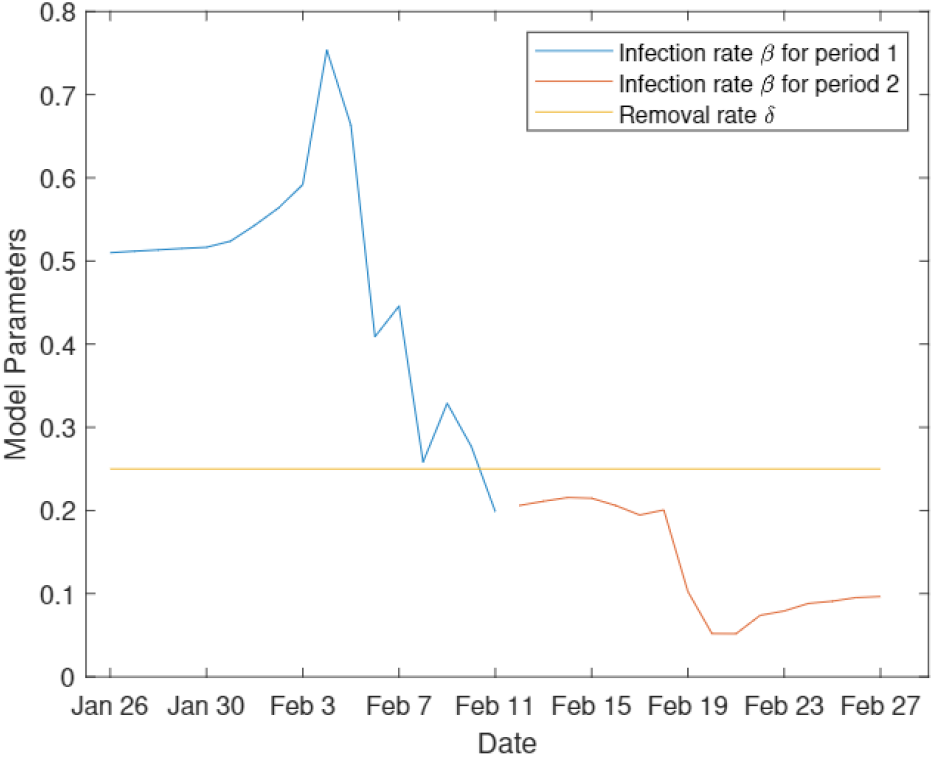
Estimated infection rate.

### 3.2 Long-term analysis with the Markovian process

To reconstruct the epidemics in Hubei province, we continuously tune the infection rate parameter *β*′ to minimize the error between the number of nodes in the removed compartment from GEMF simulations and the number of cumulative confirmed cases from the data, as shown in Fig. 4.

**Fig 4.**
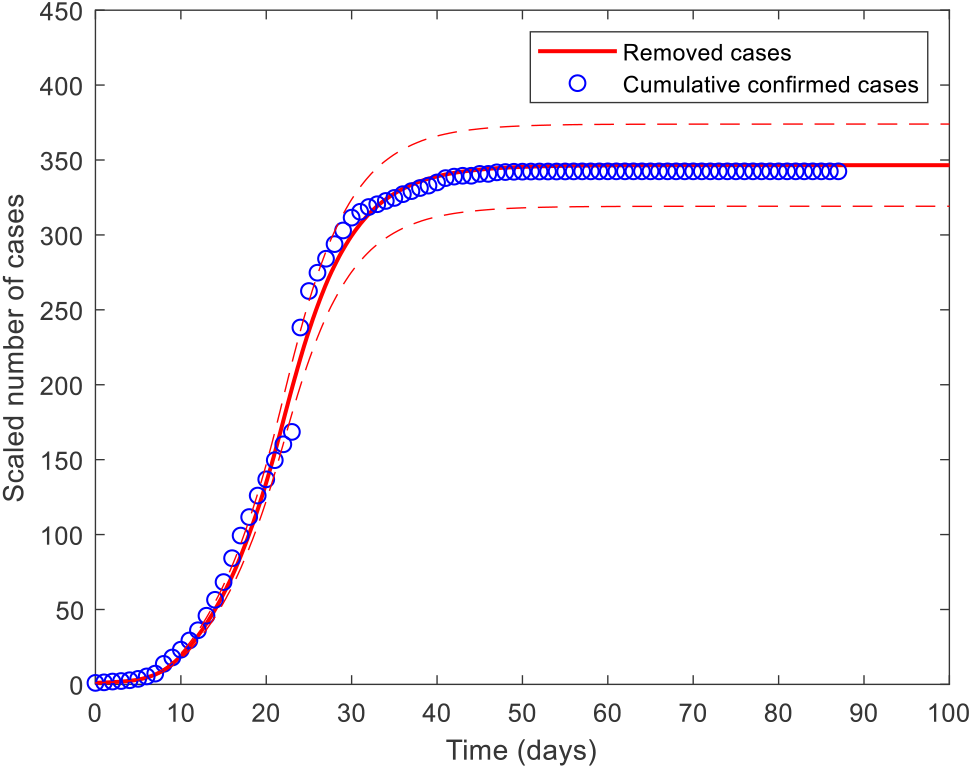
Fitting the model to the cumulative confirmed cases from January 19^th^ to April 15^th^.

By performing GEMF stochastic simulations, we obtain that *β*′ is equal to 0.06 for day 0–8 (January 19^th^–January 27^th^), and then becomes 0.0169 for day 9–21 (January 28^th^–February 9^th^), and equals 0.01 on and after day 22 (February 10^th^). The infection rate dramatically drops, indicating that public health interventions implemented in Hubei Province have considerably controlled the epidemic development since the nascent stage of the outbreak. Based on the fitted parameter *β*′, we estimate the basic reproduction number (*R*_0_ = *β*′*d*_*t*_/*δ*) decreases from 3.6 before January 27^th^, to 1.014 between January 27^th^–February 9^th^, and to 0.12 on and after February 10^th^.

Fig. 5 shows the epidemic trajectories under four scenarios, in which different mitigation strategies are implemented after February 9^th^: continuing the current trend with *β*′ = 0.0169 and the network average node degree *d*_*t*_ = 15 (Scenario 1); reducing *d*_*t*_ from 15 to 3 (Scenario 2); decreasing *β*′ by 75% (Scenario 3); reducing *β*′ from 0.0169 to 0.01 and *d*_*t*_ from 15 to 3 (Scenario 4). Preventive measures such as wearing masks can help reduce the infection rate. On the other hand, the average node degree refers to the possible contacts a person may have in daily life, and this can be reduced by contact reduction and social distancing.

**Fig 5.**
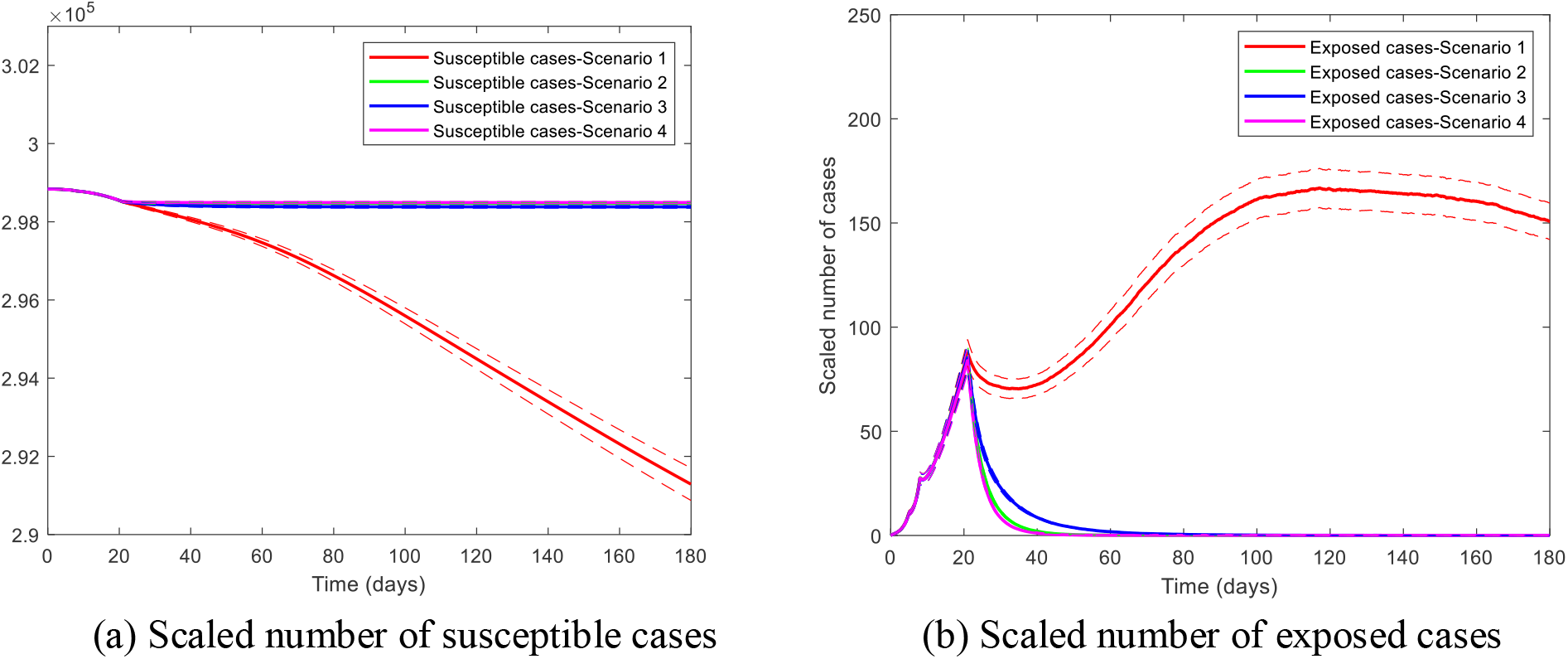
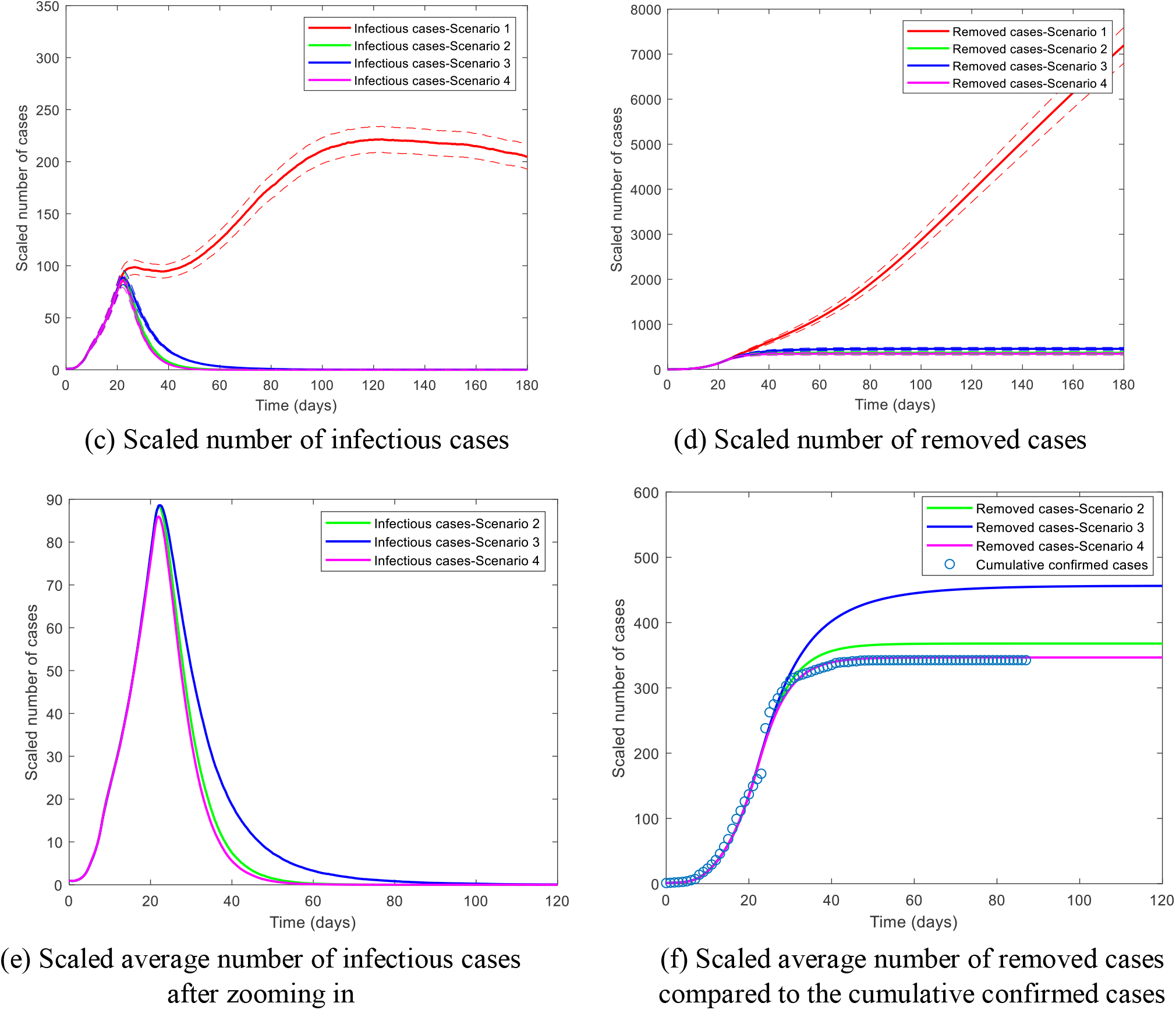
Epidemic dynamics under four scenarios (red, green, blue, and magenta) from GEMF stochastic simulations. Note that the magenta curves almost overlap perfectly with the green curves in Fig. 5(c), so Figs. 5 (e–f) are provided to zoom in and show the differences.

In Fig. 5, if the current trend as of February 9^th^ had continued, the epidemic spreading could have become more severe than what happened in Hubei Province. Reducing the average node degree from 15 to 3 (Scenario 2), the epidemic would reach the peak at around day 21.85 and fade out at around day 66.80 with the final epidemic size 367.86 (95% 339.55–396.18). When *β*′ is reduced by 75% in Scenario 3, the number of removed cases in the steady-state would reach 456.47 on average (95% CI: 423.34–489.59), and the epidemic would fade out at day 110.90 on average. When a combination of reducing *β*′ and average node degree is conducted in Scenario 4, the epidemic dynamics would approximate the actual epidemic trajectory in Hubei Province in the steady-state with mean scaled number of removed cases 346.54 (95% CI: 319.12–373.96). In addition, the epidemic would reach the peak at day 21.50 and fade out at day 61.55 on average.

Overall, GEMF simulation results suggest that more strict prevention and control measures have been taken since February 9^th^ to achieve the current cumulative confirmed cases in Hubei Province. Preventive measures and social distancing measures need to be taken in combination by the government to speed up the containment of the epidemic and reduce the peak infection dramatically.

### 3.3 Long-term analysis with the non-Markovian process

With mean infectious period (from compartments *I* → *R*) set as 3.9 days and mean incubation period (from compartments *E* → *I*) set as 5 days, we first use ordinary differential equations based on Markov processes to estimate the infection rate *β* from compartments *S* → *E*. Assuming that the epidemic starts from December 1^st^, *β* is equal to 0.68 to match the number of cumulative confirmed cases in Wuhan city until February 12^th^.

Second, we simulate the disease spread in Wuhan city with transmission between compartments incorporating non-Markovian dynamics. The transition from compartment *S* → *E* still evolves according to an exponential distribution with *β* = 0.68, while transitions from compartments *E* → *I* and *I* → *R* follow the lognormal distributions from (Linton et al., 2020; Sanche et al., 2020) in Fig. 6 (a–b). Note that the width of the infectious period becomes much shorter (blue curve) after January 18^th^, as shown in Fig. 6 (a), because the government has started to trace back and monitor those people who have contact with confirmed cases and built temporary hospitals to hospitalize both patients with mild and severe symptoms since mid-January. In Fig. 6 (a–b), we also show curves of different exponential distributions compared with these lognormal distributions for demonstration purposes.

**Fig 6.**
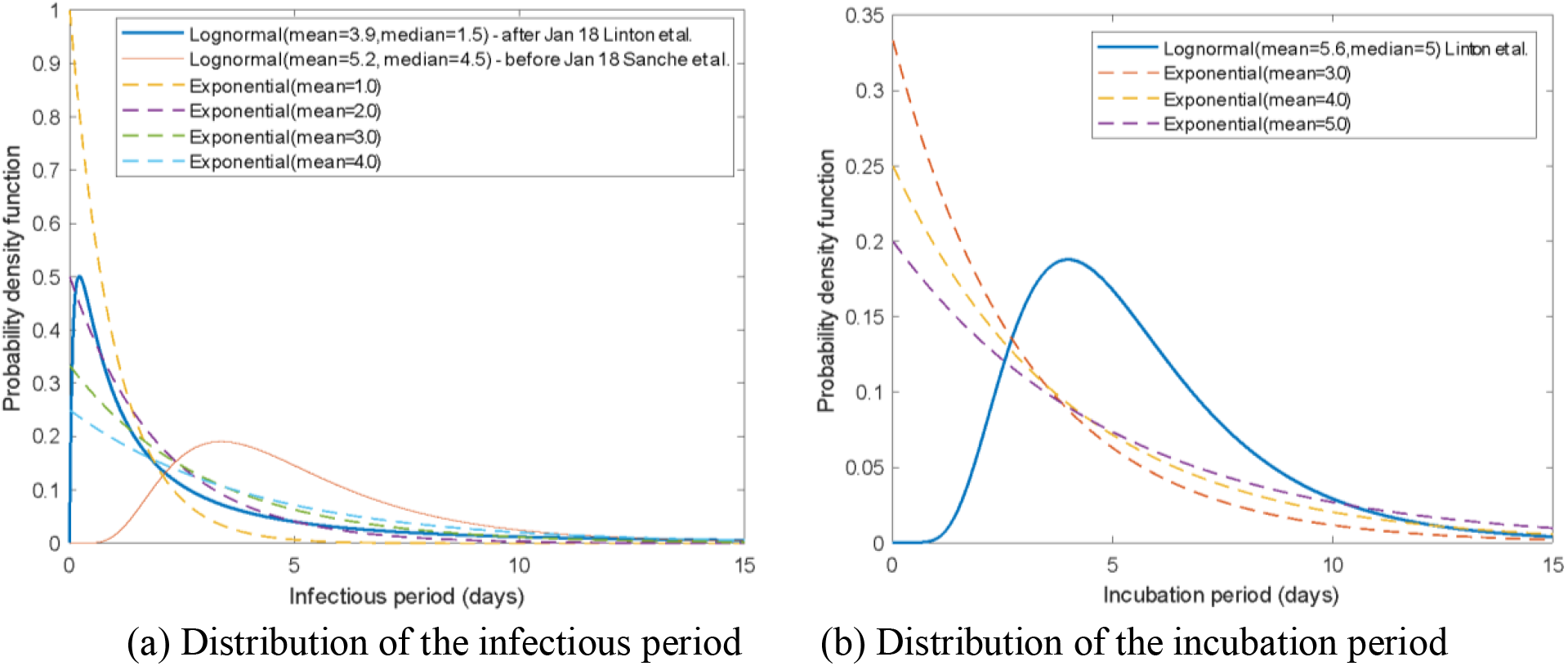
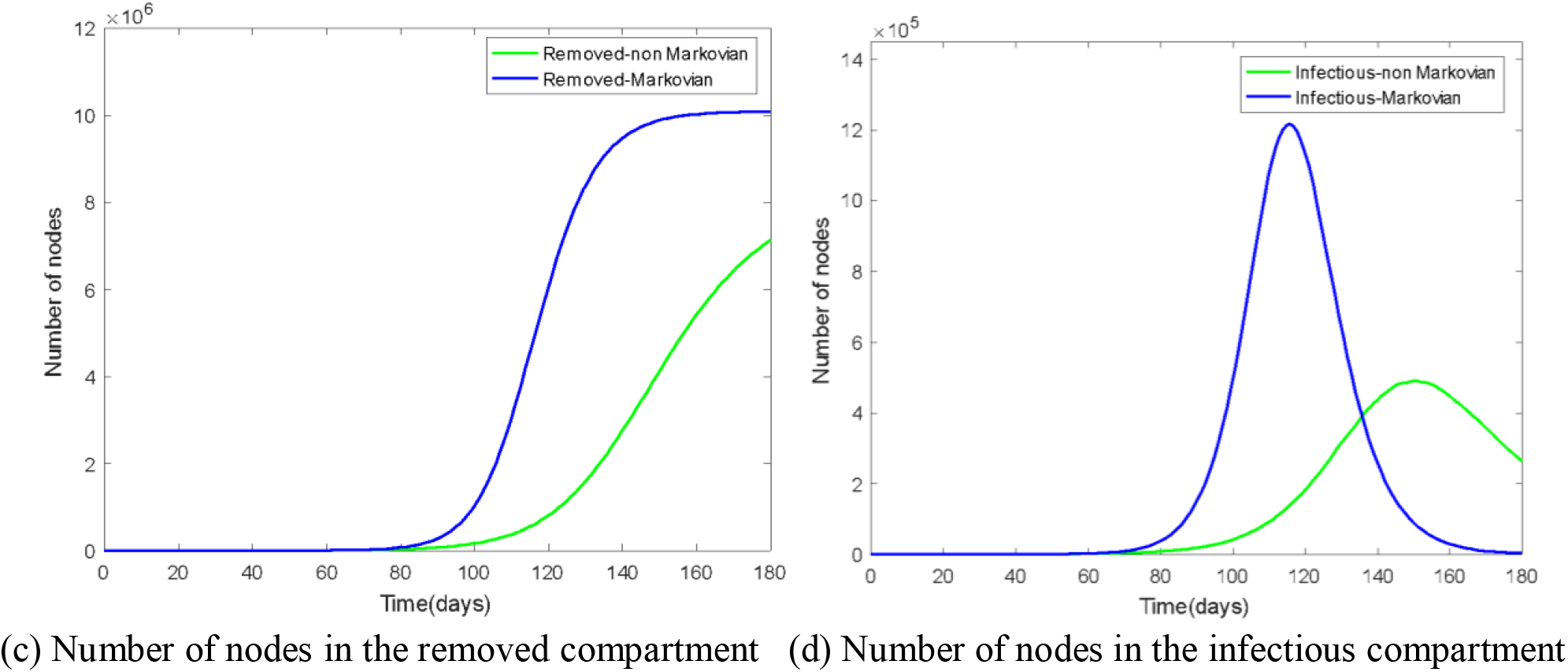
Comparison between results based Markovian process and non-Markovian process.

In Fig. 6 (c–d), simulated results incorporating non-Markovian processes deviate from the results obtained from Markov processes. More specifically, the epidemic curve with non-Markovian processes is flattened with a smaller epidemic peak compared with that from Markov processes. This is because the infectious period used in the Markovian model has an exponential distribution (mean =3.9 days) similar to the shape of the exponential distribution (mean=4.0 days) in Fig. 6 (a), but shorter than the lognormal distribution (mean=3.9 days, median=1.5 days) used after January 18^th^. The reasons above indicate that control measures to reduce the infectious period can effectively mitigate the outbreak.

On the other hand, these two methods with different distributions but with the same mean infectious period lead to inconsistent results. Since the actual infectious period and incubation period do not follow exponential distributions, these simulations suggest that future works need to consider distributions of epidemiological characteristics to better capture the COVID-19 spreading trajectory. Instead of taking mean values derived from clinical studies as input to the epidemic models, researchers need to take into account the empirically derived distribution of the parameters.

## 4. Conclusion

In this work, the ensemble Kalman filter is used to make short-term predictions of the COVID-19 cases in Wuhan City, which resulted in accurate forecasts of daily case reports. The model is able to predict daily cases and the epidemic peak. Knowing the daily cases from forecasts three days in advance allows for the proper resource allocation. The ensemble Kalman filter also allows parameter estimation, which is extremely useful for modeling purposes.

From an epidemiological modeling perspective, we simulate the epidemic spreading in Wuhan city based on non-Markovian processes, and show that different distributions with the same mean infectious period can lead to inconsistency in terms of epidemic dynamics. Since most current network-based approaches are based on Markov processes, in which epidemiological parameters are assumed to follow exponential distributions, non-Markovian process-based models need to be further explored to better predict the COVID-19 spreading. Our results show that reducing the infectious period via measures such as early case identification and isolation can reduce the epidemic size significantly.

In the long-term perspective, we test and confirm the effectiveness of non-pharmaceutical interventions on the containment of the epidemic dynamics by performing GEMF stochastic simulations. The daily cases of the COVID-19 epidemic in Hubei Province peaked at over 15,000 on February 12^th^ and became almost zero in mid-March (Xinhua, 2020). Our simulation results indicate that the containment of this highly contagious disease was achieved by stringent control measures by the Chinese government. Without continued and aggressive control measures, the epidemic in Hubei Province would have become more severe. Only with combined implementation of enhanced protective measures and social distancing measures, the epidemic dynamics would peak at around mid-February and approximate the actual epidemic trajectory in March. This can be an important message for countries going through the exponential growth of the epidemic in the current days.

## Data Availability

All data generated or analysed during this study are included in this manuscript.

## Acknowledgement

This work was supported by the Department of the Army, U.S. Army Contracting Command, Aberdeen Proving Ground, Natick Contracting Division, Ft Detrick, MD (DWFP grant W911QY-19-1-0004) and the National Science Foundation under Grant Award IIS-2027336. Any opinions, findings, and conclusions or recommendations expressed in this material are those of the authors and do not necessarily reflect the position or the policy of the Government and no official endorsement should be inferred.

## Appendix

### A. Method

#### A1. Data collection

The population size of each city in Hubei Province is obtained from the Hubei Statistical Yearbook (Hubei Provincial Bureau of Statistics, 2020). We collected Baidu Migration data (https://qianxi.baidu.com/) for the 17 cities in the Hubei Province from January 1^st^ to January 29^th^, 2020. The daily Baidu migration data contain two types of data. First type: the data include the immigration index and emigration index of each city, which are linearly related to the human traffic volume moving in and out of individual cities. Second type: for each city *i*, the data also contain the composition of city *i*’s incoming flow, namely the fraction of flows coming from every other city *j* to city *i* (flow from city *j* to *i* divided by the total flow coming into city *i*). Similarly, the data contain the fraction of flows going out of city *i* to city *m* (flow from city *i* to *m* divided by the total flow going out of city *i*).

Based on the fact that around 5 million people have moved out of Wuhan before the city lockdown on January 23rd (Zhu et al., 2020), we obtain the number of people that traveled between the cities using the following approach. First, we estimate value of *α*, which equals 5 million divided by the sum of Wuhan’s emigration indexes from January 1^st^ to January 23^rd^. Then, we can calculate the population flow moving in or out of each individual city by *α* times the immigration or emigration index. In addition, the population flow multiplied by the flow fraction going to or coming from a specific city gives us the number of daily travelers to or from a particular individual city to another.

## Notes

### Competing Interest Statement

The authors have declared no competing interest.

